# Specialty palliative care use among cancer patients: A population-based study

**DOI:** 10.1101/2024.10.31.24316547

**Authors:** J. Brian Cassel, Donna McClish, David Buxton, Leanne Yanni, Seth Roberts, Nevena Skoro, Peter May, Egidio Del Fabbro, Danielle Noreika

## Abstract

**Background:** Rigorous population-based assessments of the use of specialty palliative care (SPC) in the US are rare.

**Settings/subjects:** This study examined SPC use among cancer patients in a mid-sized metropolitan area in Southeast US.

**Measurements:** In this cancer decedent cohort study, data were acquired and linked from the state-wide cancer registry; state-wide hospital discharge dataset; and local SPC providers.

**Results:** 12,030 individuals with cancer were included in this study; only 2,958 (24.6%) used SPC. Of the 9,072 persons who did not use SPC, 3,877 (42.7%) went only to hospitals that did not offer SPC; and 3,517 (38.8%) went to hospitals that offered SPC but did not use it. About half of SPC recipients (1493; 50.5%) first received SPC in the final 30 days of life, including 768 (26.0%) in the final week of life. Characteristics associated with using SPC use included being in an socio-economic status quintile other than the lowest; being younger; being Black; having a solid (versus hematological) cancer; having a shorter survival with cancer; dying in the latter two years of the study; being from an area of low or complete rurality; having a hospital admission in the final 60 days prior to initiation of PC or death; having more days in hospital; and living within 15 miles of a hospital offering SPC.

**Conclusions:** In this population-based study, only one-quarter of cancer patients used SPC, and for half who did so, it came in the final 30 days of life.

**Key Message:** This novel population-based study found that about 25% of cancer patients who died 2012-2015 had used of specialty palliative care (SPC). Half of them first received SPC in the final month of life. Characteristics associated with SPC included being Black, younger, and residing within 15 miles of a hospital offering SPC.

## Background

American Society of Clinical Oncology (ASCO) guidelines recommend the integration of specialist palliative care (SPC) into oncology care(1). Reviews of randomized trials found that SPC improves patient and family experience, including symptom burden and psychological distress, and increased probability of death in preferred place(2,3).

While SPC capacity has increased substantially over the last two decades in the US(4–7) and internationally(8,9), most people with palliative care needs do not receive SPC(10,11). In some cases this may reflect patient preferences(12), but other key determinants include insufficient PC capacity, sub-optimal referral rates among oncologists and low awareness of PC among patients(13–15). There are potential inequities in access to scarce SPC capacity, including by diagnosis, race and ethnicity, socioeconomic position, and urban and rural locations(16–18).

There is a need for better evidence to understand health disparities US(19,20). Robust data on how many people with cancer in the US receive SPC, and the characteristics and representativeness of that population, are rare for several reasons. First, people with cancer receive care across different settings without routine data linkage(10). Second, it is often difficult to determine which people actually received care from palliative care specialists using a single source of secondary data(21).

### Rationale and aims

In the context of these evidence needs and data challenges, we compiled a population-representative dataset to understand SPC engagement among people with cancer in the Richmond-Petersburg metropolitan statistical area (Richmond MSA) in Virginia for 2010-2015. Our specific research questions were: (1) what proportion of people with cancer in the Richmond MSA used SPC? (2) What socio-demographic and clinical characteristics were associated with SPC receipt?

## Methods

This was a population-based decedent cohort study using secondary data. Data sources were the statewide cancer registry; statewide hospital discharge data; and medical groups’ specialty palliative care encounters. The Richmond MSA includes 14 cities and counties and had 1.3 million people at the time of the study(22). The population characteristics of the population in 2011-2015(23) are shown in Table 1 for the Richmond MSA and the nation. Richmond MSA characteristics are similar to the nation, though with greater percentage of Black persons, greater percentage of English-only speakers, and higher cancer mortality-incidence rate.

**Table 1.** Richmond MSA and national population characteristics 2011-2015.

| Demographic, social, and economic characteristics | Richmond MSA<br>Measure and<br>margin of error | National<br>Measure and<br>margin of error |
| --- | --- | --- |
| Age |  |  |
| 18 years and over | 77.5% (0.1%) | 78.3% (0.1%) |
| 65 years and over | 13.3% (0.1%) | 17.7% (0.1%) |
| Race |  |  |
| White (only) | 61.9% (0.1%) | 60.5% (0.1%) |
| Black / African American (only) | 30.0% (0.1%) | 12.1% (0.1%) |
| American Indian (only) | 0.4% (0.1%) | 1.0% (0.1%) |
| Asian (only) | 3.6% (0.1%) | 6.0% (0.1%) |
| Native Hawaiian / other Pacific Islander (only) | 0.0% (0.1%) | 0.2% (0.1%) |
| Other race (only) | 1.4% (0.1%) | 7.4% (0.1%) |
| Two or more races | 2.6% (0.1%) | 12.8% (0.1%) |
| Hispanic or Latino (of any race) | 5.5% (0.1%) | 19.4% (0.1%) |
| Education |  |  |
| <9<sup>th</sup> grade | 4.3% (0.2%) | 4.6% (0.1%) |
| 9-12<sup>th</sup> grade no diploma | 7.6% (0.2%) | 5.6% (0.1%) |
| High school graduate including equivalency | 26.4% (0.4%) | 25.9% (0.1%) |
| Some college, no degree | 21.1% (0.4%) | 18.9% (0.1%) |
| Associate's degree | 6.9% (0.2%) | 8.8% (0.1%) |
| Bachelor's degree | 21.1% (0.3%) | 21.8% (0.1%) |
| Graduate or professional degree | 12.6% (0.2%) | 14.3% (0.1%) |
| Language spoken at home |  |  |
| English only | 90.1% (0.2%) | 77.5% (0.1%) |
| Language other than English | 9.9% (0.2%) | 22.5% (0.1%) |
| Speak English less than "very well" | 3.9% (0.1%) | 8.7% (0.1%) |
| Economic characteristics |  |  |
| Unemployment rate in civilian labor force | 7.5% (0.3%) | 4.3% (0.1%) |
| Median household income | \$59,919 (\$647) | \$77,719 (\$186) |
| With food stamp /SNAP benefits past 12 months | 10.8% (0.3%) | 12.2% (0.1%) |
| Families with income in past 12 months below<br>poverty level | 8.8% (0.3%) | 8.8% (0.1%) |
| Cancer mortality-incidence age-adjusted rate (per<br>100,000) | 463.16 | 449.21 |

Sources: US Census American Community Survey data(23); US Centers for Disease Control WONDER(24).

### Data Sources

The Virginia Cancer Registry (VCR) provided statewide data on May 7, 2018 on adult cancer decedents (>= 21 at time of death) who died 2012-2015. Data elements from the VCR included patient-level identifiers and demographics; and tumor-level diagnosis dates, cancer types, histology, stage at diagnosis, and locality of residence (including census tract) at time of diagnosis.

Virginia Health Information (VHI) provided statewide data on all hospitalizations from 2010-2015 for those cancer decedents on September 12, 2019. Data elements included admission and discharge dates, diagnoses, procedures, charges, insurance type, and hospital identifiers. The three palliative care programs provided patient-level data on SPC services from 2010-2015 in December 2019. Data elements included patient identifiers and dates of services.

Authors representing the three palliative care providers (D. Buxton, D. Noreika, E. Del Fabbro, L. Yanni, S. Roberts had access to patient identifiers only for their own programs prior to submitting their data. One author (N. Skoro) had access to patient identifiers in the data from the cancer registry and palliative care provider groups; the data were de-identified prior to further analysis.

### Availability of SPC

Information about availability of SPC services was obtained directly from the SPC programs. Within the Richmond MSA, there were 3 distinct medical groups providing SPC services at 6 hospitals. One was an academic medical center providing hospital-PC and clinic-PC; a second group was at Catholic, non-profit system with four hospital locations offering hospital-PC, office-PC and home-PC; and a third group was at a for-profit hospital. All three programs had multi-disciplinary teams; two were certified by the Joint Commission’s hospital-PC certification program(25). These were the only SPC providers in the Richmond MSA in the study time period. Emails, websites, and phone calls confirmed that other hospitals within or nearby the area did not have SPC services during the years of interest (2010–2015), and that no other medical groups or insurance-based providers were operating in the region during that time. The two three hospitals that did not offer SPC were for-profit. None of the 5 small, rural hospitals just outside of the MSA offered palliative care services at that time. See Figure 1.

**Figure 1.**
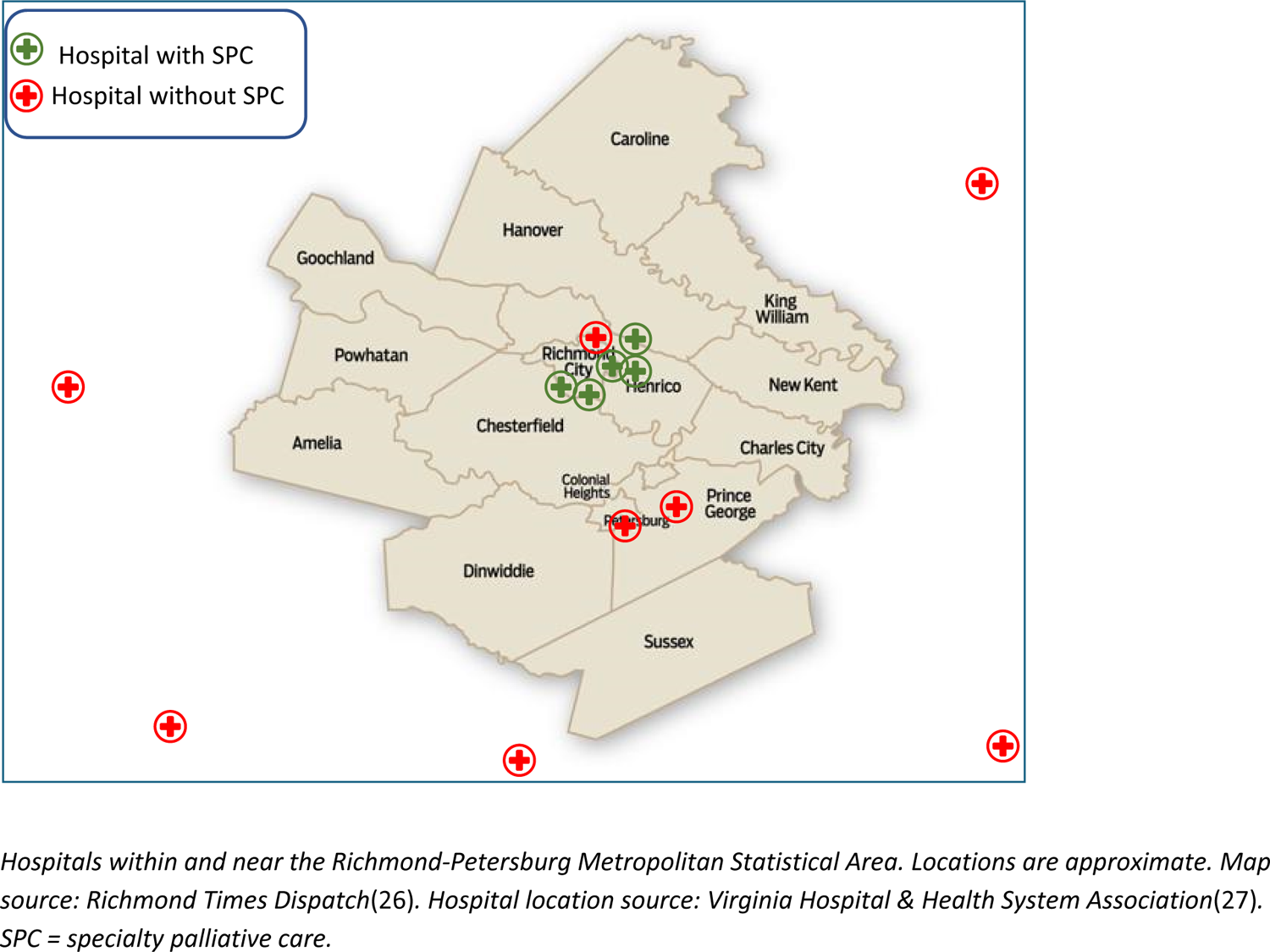
Availability of Specialist Palliative Care within and around the Richmond MSA

### Data linkage

VCR data were linked to hospitalization data by VHI using hashed (encrypted) social security numbers (SSN). VCR data were linked by the researchers to SPC data using patient name and date of birth. The resulting VCR-VHI-SPC dataset was then linked to US census data based on census tract identifiers at the time of diagnosis.

### Inclusion and Exclusion Criteria

VCR data included cancer registry cases (primary malignant neoplasms) for people who were at least 21 years old at the time of diagnosis and were known to have died between 2012-2015. VCR excluded cases that were diagnosed or treated within the Veterans Health Administration system per VCR requirements, and we further excluded people whose only cancer was a non-melanoma skin cancer. Using the combined VCR-VHI data, we included people that were living within the Richmond MSA at the time of cancer diagnosis, and excluded cases that had additional cancers diagnosed outside the MSA. We excluded cases that had hospitalizations outside of the Richmond MSA (beyond the several small, rural hospitals just outside the MSA, none of which had SPC services) because we had palliative care data only from providers within the MSA.

In the dataset provided by the VCR there were 13,260 adult cancer decedents who died at age 21+ in years 2012-2015 who were living in the Richmond MSA at the time cancer diagnosis. We excluded 1,230 people because they either did not have a valid SSN (n=91) which would have prevented linking them to VHI hospitalization data; or had hospitalizations beyond the 13 hospitals within or nearby the Richmond MSA within 2 years of death (n=792) which would have prevented our assessment of their PC access or exposure; or had irreconcilable dates from multiple data sources (n=377) such as admission, discharge dates or initial PC dates after date of death, or incomplete diagnosis dates. The remaining dataset contained 12,030 cancer decedents.

### Variables used in this study

For each patient, the data included at least two years of potential hospitalizations and SPC encounters prior to death. Because we were examining factors related to receiving palliative care, the number of hospitalizations as a potential predictor variable was measured prior to initiation of SPC if received, else prior to death.

For the descriptive analyses of what proportion of the cancer population used SPC, our outcome variables included a binary variable of SPC use, and the timing of first SPC encounter prior to death. Predictor variables included age at time of cancer diagnosis, age at time of death, sex, race, cancer type, rurality, and socio-economic status (SES). A rurality index was used, based on the percent of the population living in non-urban areas divided into 4 groups representing 0% rural, 1-50% rural, 51-99% rural, and 100% rural(28). The socio-economic status variable was derived from 7 data elements from the American Community Survey census data and converted to quintiles(29). Imputation for SES was performed where only 5 or 6 of those data elements were available; quintiles were ranked from low SES to high SES. Cancers were characterized in two ways: solid versus hematologic, and poor prognosis or not. Cancers with poor prognosis were those that nationally had 40% or fewer surviving for 5 years(30,31); or had distant metastases at diagnosis.

### Statistical analyses

Frequencies and percentages were used to describe categorical variables, with means (standard deviations) and/or median (interquartile range) used for continuous variables. Univariate association with SPC used Chi-square (categorical variables), or Wilcoxon Rank Sum test (continuous variables). Multivariable analysis of SPC use used backward logistic regression, with variables staying in the model as long as p<=0.05. Cox proportional hazards regression was used to evaluate characteristics associated with timing of SPC for those who used SPC. Analyses were conducted in SAS(32).

### IRB, privacy, funding, and conflicts of interest

The Virginia Department of Health approved the study on May 2, 2018 and the VCU IRB exempted this study on January 8, 2019. Data use agreements were established with VCR, VHI, and the two medical groups outside of VCU. The privacy officers of each medical group approved the acquisition of data on SPC use. All entities waived the need for consent as this study concerned decedents. The study was funded by a pilot grant from the VCU Department of Internal Medicine and a pilot grant from the American Cancer Society; neither entity had a further role in the research or the reporting of results. The study authors report no conflicts of interest.

## Results

### Patient characteristics

Clinical, demographic, geographic and socio-economic characteristics of the cancer population are provided in the first column of Table 2. The median distance from patient residence to nearest hospital offering SPC was 5.5 miles. One-fifth (20.6%) lived in an area that was at least 51% rural. About half (55.2%) spent more than 5 days in a hospital in the last six months of life. Median age was 68 at diagnosis and 74 at death; 47.8% were female; 29.8% were Black. 11.4% had a hematologic malignancy and 40.6% had a poor prognosis cancer at time of diagnosis.

**Table 2.** Univariate analyses of SPC use (n=12,030)

| Characteristics | Total | Used SPC | Did not use SPC | p-value |
| --- | --- | --- | --- | --- |
| Age at death (mean (SD), median (IQR)) | 73.3 (13.2)<br>74.0 (20.0) | 69.4 (13.8)<br>70.0 (20.0) | 74.5 (12.8)<br>76.0 (18.0) | <0.0001 |
| Age at diagnosis (mean (SD), median (IQR)) * | 67.9 (13.0)<br>68.0 (17.0) | 64.6 (13.5)<br>65.0 (18.0) | 68.9 (12.6)<br>69.0 (17.0) | <0.0001 |
| Years from Diagnosis to Death (mean (SD), median (IQR)) * | 5.3 (7.0)<br>2.0 (7.7) | 4.5 (6.5)<br>1.4 (5.9) | 5.6 (8.3)<br>2.2 (8.3) | 0.0021 |
| Miles to hospital offering SPC (mean(SD), median (IQR)) ** | 9.3 (10.8)<br>5.5 (11.9) | 7.4 (8.0)<br>4.7 (8.9) | 9.9 (11.5)<br>5.6 (14.5) | <0.0001 |
| Hospital days 6 month prior to SPC or death (n (%)) |  |  |  | <0.0001 |
| 0 | 3025 | 326 (11.8) | 2699 (88.2) |  |
| 1-5 | 2368 | 548 (23.1) | 1820 (76.9) |  |
| >5 | 6637 | 2084 (31.4) | 4553 (68.6) |  |
| Cancer Site (first if multiple) (n (%)) * |  |  |  | <0.0001 |
| Breast | 1433 | 369 (25.7) | 1064 (74.3) |  |
| Prostate | 1545 | 295 (19.1) | 1250 (81.9) |  |
| Lung | 2533 | 597 (23.6) | 1936 (76.4) |  |
| Colo-rectal | 1240 | 316 (25.5) | 924 (74.5) |  |
| Other solid | 4093 | 1112 (27.2) | 2981 (72.8) |  |
| Hematologic | 1186 | 269 (22.7) | 917 (77.3) |  |
| Race (n (%)) |  |  |  | <0.0001 |
| White | 8248 | 1841 (22.3) | 6407 (77.7) |  |
| Black | 3581 | 1060 (29.6) | 2521 (70.4) |  |
| Other | 201 | 57 (28.4) | 144 (71.6) |  |
| Sex (n (%)) |  |  |  | 0.0141 |
| Male | 6279 | 1486 (23.7) | 4893 (76.3) |  |
| Female | 5751 | 1472 (25.6) | 4279 (74.4) |  |
| Year of Death (n (%)) |  |  |  | <0.0001 |
| 2012 | 2380 | 535 (22.5) | 1845 (77.5) |  |
| 2013 | 3203 | 701 (21.9) | 2502 (78.1) |  |
| 2014 | 3143 | 784 (24.9) | 2359 (75.1) |  |
| 2015 | 2204 | 938 (28.4) | 2366 (71.6) |  |
| Multiple Cancers (n (%)) |  |  |  | 0.1701 |
| Yes | 1906 | 475 (23.3) | 1461 (76.7) |  |
| No | 10124 | 2483 (24.8) | 7611 (75.2) |  |
| Any hematologic cancer (n (%)) |  |  |  | 0.0734 |
| Yes | 1370 | 310 (22.6) | 1060 (77.4) |  |
| No | 10660 | 2648 (24.8) | 8012 (75.2) |  |
| SES rank** (n (%)) |  |  |  | 0.6857 |
| 1<sup>st</sup> quintile | 2348 | 563 (24.0) | 1785 (76.0) |  |
| 2<sup>nd</sup> quintile | 2390 | 602 (25.2) | 1788 (74.8) |  |
| 3<sup>rd</sup> quintile | 2379 | 573 (24.1) | 1806 (75.9) |  |
| 4<sup>th</sup> quintile | 2349 | 592 (25.2) | 1757 (74.8) |  |
| 5<sup>th</sup> quintile | 2387 | 605 (25.3) | 1782 (74.7) |  |
| Rurality Group** (n (%)) |  |  |  | 0.0033 |
| 0% rural | 7543 | 1854 (24.6) | 5689 (75.4) |  |
| 1-50% rural | 1868 | 482 (25.8) | 1386 (74.2) |  |
| 51-99% rural | 678 | 133 (19.6 ) | 545 (80.4) |  |
| 100% rural | 1797 | 476 (26.5) | 1321 (73.5) |  |
| Poor Prognosis Cancer (n (%)) |  |  |  | 0.0003 |
| Yes | 4889 | 1287 (26.3) | 3602 (73.7) |  |
| No | 7141 | 1671 (23.4) | 5407 (77.6) |  |

SPC = specialty palliative care; SD = standard deviation; IQR = interquartile range; SES = socioeconomic status. \*using initial cancer if patient had multiple cancers. \*\*missing 177 for SES; missing 144 for rurality; missing 199 for zip code which was used for computing miles from hospitals offering SPC.

### Use of local hospitals

Hospital admission was evaluated at 24, 18 and 6 months before death. Looking at all 12,030 cancer decedents and the full 24-month period, 10,273 (85.4%) went to a hospital; of them, 8,051 (78.4%) went to a hospital offering SPC. In the 18 months before death, 9,914 (82.4%) had at least one hospitalization, and 7,671 of them (77.4%) went to a hospital offering SPC. In the 6 months before death, 9005 (74.9%) were hospitalized at least once and 6,815 of them (75.7%) went to a hospital offering SPC.

Some patients used more than 1 hospital. Of the 6,614 patients who had at least 1 hospitalization the 30 days prior to PC or death, 344 (5.2%) went to more than 1 hospital. Of the 8,301 patients who had at least 1 hospitalization over the 3 months prior to PC or death, 921 (11.1)% went to more than 1 hospital. Of the 9,005 patients who had at least 1 hospitalization over the 6 months prior to PC or death, 1,333 (14.8%) went to more than 1 hospital.

### Use and timing of specialty palliative care

One-quarter of the cancer decedents (n=2,958, 24.6%) received SPC from one or more sources in the 18 months before death. Of the 7,671 who went to a hospital offering SPC within 18 months before death, 2,671 (34.8%) received SPC; of the other 4,359 who did not go to a hospital offering SPC, 287 (6.6%) received SPC in clinic or home settings. While most SPC recipients (97.5%) received SPC from a single medical group, 73 received SPC from two medical groups and 1 person received SPC from all 3.

Of the 9,072 persons who did not use palliative care, 3,877 (42.7%) went only to hospitals that did not offer SPC; 3,517 (38.8%) went to hospitals that offered SPC but did not use it; and the remaining 1,678 (18.5%) did not go to any hospital (and did not use community-based SPC).

From first cancer diagnosis to first SPC encounter, a mean of 4.5 years passed (SD 6.5), with a median of 1.4 years (IQR 5.9). From first SPC encounter to death, the mean was 123.9 days (SD 243) with a median of 30 days (IQR 107). From first SPC encounter to death, 1493 (50.5%) were in the final 30 days of life, including 768 (26.0%) in the final week of life, 450 (15.2%) in the final 3 days of life, and 126 (4.3%) had a first SPC encounter on the day they died.

### Characteristics associated with SPC use and timing

Most of the characteristics of the population that we evaluated were significantly associated with whether SPC was used, as shown in Table 2. The rate of SPC use increased over the years, from around 22% for those dying in 2012 or 2013, and increasing to 28% by 2015.

As shown in Table 3, the following characteristics were associated with using SPC in the multivariate analysis: being in an SES quintile other than the lowest; being younger; being Black; having a solid (versus hematological) cancer; having a shorter survival with cancer; dying in the latter two years of the study; being from an area of low or complete rurality; having a hospital admission in the final 60 days prior to initiation of PC or death; having more days in hospital; and living within 15 miles of a hospital offering SPC. The AUC statistic was 0.749.

**Table 3.** Multivariate analysis of SPC use (n=11,689)

| Characteristics | OR (95% CI) |
| --- | --- |
| SES |  |
| Rank 2 vs 1 | 1.27 (1.09, 1.49) |
| Rank 3 vs 1 | 1.30 (1.11, 1.53) |
| Rank 4 vs 1 | 1.41 (1.20, 1.66) |
| Rank 5 vs 1 | 1.53 (1.30, 1.80) |
| Age at death (10-year increments) | 0.78 (0.75,0.80) |
| Black | 1.37 (1.26, 1.57) |
| Any hematological malignancy | 0.81 (0.70, 0.93) |
| Less than 15 miles to hospital offering SPC | 2.66 (2.33, 3.04) |
| Years from diagnosis to death |  |
| <1 month vs >1 year | 1.80 (1.58, 2.05) |
| 1-2 months vs >1 year | 1.11 (0.90, 1.36) |
| 2-12 months vs > 1 year | 0.98 (0.87, 1.10) |
| Death year |  |
| 2013 vs 2012 | 1.12 (0.98, 1.28) |
| 2014 vs 2012 | 1.29 (1.13, 1.48) |
| 2015 vs 2012 | 1.60 (1.40, 1.83) |
| Rurality group |  |
| 1-50% vs 0% rural | 1.23 (1.08, 1.39) |
| 51-99% vs 0% rural | 1.02 (0.82, 1.27) |
| 100% vs 0 % rural | 1.77 (1.53, 2.04) |
| Hospital days in 6 months prior to SPC or death |  |
| 1-5 vs 0 days | 3.79 (3.18, 4.53) |
| >5 vs 0 days | 5.81 (4.96, 6.80) |

SPC = specialty palliative care; SES = socioeconomic status; OR = odds ratio; CI = confidence interval.

Two secondary regressions were performed: one model limited to patients admitted to hospitals offering SPC (to control for access to SPC), and one model limited patients with poor-prognosis cancers or stage (which are commonly targeted in SPC intervention studies). Similar AUCs were achieved and certain variables were significant predictors across models: younger age, being Black, more recent year, shorter distance to SPC hospital, rurality group, and more days in hospital in final 6 months. See Supporting Information. Univariate analyses and Cox proportional hazards regression were also conducted to determine characteristics associated with the timing of SPC relative to death, for those who used SPC (n=2,792).

Few variables were statistically significant in the multivariate analysis, as shown in Table 4. The characteristic associated with longest time from SPC to death was having zero hospitalizations in the six months prior. A number of variables that were associated with SPC use, such as race, cancer type, rurality and SES, were not significantly associated with the timeliness of SPC among those who used it.

**Table 4.** Characteristics associated with fewer days from SPC to death, multivariate analysis.

| Characteristics | N | Mean days | Median days | HR (95% CI) |
| --- | --- | --- | --- | --- |
| Age |  |  |  |  |
| Age <50 | 223 | 101.4 | 43 | -- |
| Age >50 | 2569 | 72.3 | 25 | 1.31 (1.14, 1.51) |
| Death year |  |  |  |  |
| 2012 | 524 | 54.8 | 19 | -- |
| 2013 | 675 | 74.1 | 22 | 0.858 (0.76, 0.96) |
| 2014 | 733 | 75.6 | 29 | 0.842 (0.75, 0.94) |
| 2015 | 860 | 86.1 | 34 | 0.784 (0.70, 0.88) |
| Time from diagnosis to SPC |  |  |  |  |
| 0-1 month | 517 | 58.4 | 16 | -- |
| 1-2 months | 156 | 68.0 | 27 | 0.86 (0.72, 1.04) |
| >2 months | 2119 | 79.0 | 29 | 0.79 (0.72, 0.88) |
| Number of hospitalizations 6 months prior |  |  |  |  |
| 0 | 198 | 137.3 | 90 | -- |
| 1 | 1173 | 79.7 | 28 | 1.44 (1.23, 1.68) |
| 2 | 731 | 65.3 | 23 | 1.68 (1.43, 1.97) |
| >2 | 690 | 57.7 | 17 | 1.85 (1.57, 2.17) |

SPC = specialty palliative care; HR = hazard ratio; CI = confidence interval.

## Discussion

In this population-based study of specialty palliative care use by cancer patients in a mid-sized metropolitan area in the US, only one quarter of the cancer population (24.6%) received SPC, and this often came near the end of life, with 50% first receiving SPC in the final 30 days of life. There was some improvement over the years of the study, with 22% of those who died in 2012 or 2013 receiving SPC, 25% of those of who died in 2014, and 28% of those who died in 2015.

By today’s standards, this is suboptimal cancer care. It is unclear why SPC was “too little, too late” for cancer patients. The answer does not seem to be access alone. Most (6 of 8) hospitals within the MSA offered SPC. Most patients in the study (82.4%) had a hospital admission in the 18 months before death, and most of those (77.4%) went to a hospital offering SPC. While distance to the closest hospital offering SPC is a significant predictor of SPC use, the median distance for SPC users (7.4 miles) and non-SPC users (9.9 miles) is similar.

One could hypothesize that the low uptake is due to the culture of cancer care and the visibility of SPC in local cancer centers. At the time of the study (deaths occurred 2012-2015), the hospitals providing SPC had mature programs with multi-disciplinary teams. Bakitas’s “Project Enable II” trial had been published in 2009(33); Temel’s landmark trial had been published in 2010(34); the Joint Commission’s certification of hospital-PC programs was launched in 2011(25); and ASCO’s provisional clinical guidelines for palliative care were published in 2012(35). Yet the number of non-users of SPC who went to a hospital offering SPC (3,517) was almost as large as those who went to hospitals lacking SPC (3,877).

Significant variation in SPC use was found; 9 variables were associated with SPC use including socio-economic status; younger age; race (being Black); cancer type (solid tumors); length of survival with cancer; rurality; hospital use; and shorter distance to a hospital offering SPC. Fewer variables were Fewer variables were associated with the timing of SPC relative to death, among those who used SPC. This points to multiple potential sources of inequity. Two noteworthy findings were that SPC use was greater among younger patients and Black patients.

In this study, Black cancer patients were more likely to use SPC than White cancer patients, 29.6% versus 22.3%. This is not attributable to a difference in the hospitals they went to; of those hospitalized in the last six months of life, Black patients and other patients were about equally likely (76.8% versus 75.2%) to go to a hospital offering SPC. Many studies of end-of-life healthcare in the US have documented lower use of hospice by Black compared to White patients; for example, 35.6% of Black Medicare decedents versus 50.5% of White Medicare decedents had enrolled in hospice in 2023(36). Research on race differences in non-hospice SPC is less common, and much of the research on the prevalence of SPC has relied upon invalid measures of SPC use(21).

Other studies of palliative care prevalence for cancer patients have been conducted; their methods and definitions of palliative care differ substantially. A survey-based study in Japan estimated a 24% SPC rate for cancer patients(37). A study in Belgium estimated 56.4% SPC use for hematological cancers, and 73% for most solid cancer types(38). Several studies in Ontario have estimated SPC prevalence and its impact for cancer patients(39–42), with one finding 75.6% of hospitalized patients with terminal illnesses such as cancer accessed SPC(40). Most studies in the US purporting to evaluate population-level prevalence of palliative care services have serious methodological flaws(21). Two rigorous studies found lower rates than the current study. Hugar et al. evaluated SPC among patients with bladder cancer 2008-2013, finding that 3.6% received SPC(43); and Hua et al. evaluated patients with metastatic cancer 2017-2019, finding 12.0% received SPC(44). Those two studies used national Medicare data; the current study employed a geographically focused dataset inclusive of all insurance types and all ages.

### Methodological strengths

The current study used population-based data sets appropriate for determining population-level use of SPC. The starting place was population data in the form of the state cancer registry. This was then linked to another population data source, state-wide hospital discharges. Finally, institutional data on SPC use was acquired from SPC providers in the MSA.

This approach had several strengths. Not all patients were hospitalized, and because early palliative care may reduce the need for subsequent hospitalizations(45–47), it was critical to start with all cancer decedents regardless of hospital use. Second, we used statewide cancer registry and hospital discharge data to exclude people who were diagnosed or hospitalized outside of the metropolitan area, thus focusing the analysis on people for whom we could evaluate SPC use. Most importantly, we did not rely on the ICD9/10 codes for “palliative care encounter” (V66.7 / Z51.5) which are not suitable for research on SPC(21).

To our knowledge, this approach has not been used in prior studies of SPC in the US. It is a replicable method that would work well in other small- and medium-sized metropolitan statistical areas. The challenge of using this approach in larger areas would be in securing data use agreements with larger numbers of SPC providers.

## Limitations

This study was limited to a particular metropolitan area and findings may not be generalizable to other metropolitan areas or regions. The census data in Table 1 demonstrate that the population of this metropolitan area is not unusual. The study was limited to adults with cancer, and findings may not be applicable to pediatric populations or other diseases. Socio-economic characteristics were measured at the census tract level (not the patient level); this introduces some degree of measurement error.

Information on comorbidities and insurance type are available in the hospital discharge data, but the analyses in this study were not limited to those who were hospitalized. It is possible that some people received some healthcare outside of Virginia.

The study examined healthcare that occurred between 2010 and 2015, and may no longer be representative of care in the region. However, informal interviews with palliative care leaders in the region have indicated that the scope and volume of SPC for cancer patients remains largely the same. While some time has passed since the data or phenomena, no other study in the US has been published in the interim, using population-based data. Future work using similar population-based methods in other metropolitan areas, in more recent years, would address some of the limitations of this study.

## Conclusion

To our knowledge this is the first study in the US to combine state-wide population-level datasets with local SPC encounter data from SPC providers, to produce a regional evaluation of SPC use. This population-based study of specialty palliative care use among cancer patients found that 25% used SPC, identified 9 variables associated with SPC use, and found that Black patients were more likely to use SPC, in contrast with national data on hospice use.

## Data Availability

Data cannot be shared publicly because of prohibition of such in the data use agreements with the data providers.

